# Redefining the multidimensional clinical phenotypes of frontotemporal lobar degeneration syndromes

**DOI:** 10.1101/19012260

**Authors:** Alexander G. Murley, Ian Coyle-Gilchrist, Matthew Rouse, P Simon Jones, Win Li, Julie Wiggins, Claire Lansdall, Patricia Vázquez Rodríguez, Alicia Wilcox, Kamen A. Tsvetanov, Karalyn Patterson, Matthew A. Lambon Ralph, James B. Rowe

## Abstract

The syndromes caused by frontotemporal lobar degeneration (FTLD) have highly heterogenous and overlapping clinical features. There has been great progress in the refinement of clinical diagnostic criteria in the last decade, but we propose that a better understanding of aetiology, pathophysiology and symptomatic treatments can arise from a transdiagnostic approach to clinical phenotype and brain morphometry. In a cross-sectional epidemiological study, we examined 310 patients with a syndrome likely to be caused by frontotemporal lobar degeneration, including behavioural variant frontotemporal dementia (bvFTD), the non-fluent (nfvPPA), semantic (svPPA) variants of primary progressive aphasia, progressive supranuclear palsy (PSP) and corticobasal syndrome (CBS). We also included patients with logopenic primary progressive aphasia (lvPPA) and those who met criteria for PPA but not one of the three subtypes. To date, forty-nine patients have a neuropathological diagnosis. A principal component analysis identified symptom dimensions that broadly recapitulated the core features of the main clinical syndromes. However, the subject-specific scores on these dimensions showed considerable overlap across the diagnostic groups. Sixty-two percent of participants had phenotypic features that met the diagnostic criteria for more than one syndrome. Behavioural disturbance was prevalent in all groups. Forty-four percent of patients with CBS had PSP-like features and thirty percent of patients with PSP had CBS-like features. Many patients with PSP and CBS had language impairments consistent with nfvPPA while patients with bvFTD often had semantic impairments. Using multivariate source-based morphometry on a subset of patients (n=133), we identified patterns of co-varying brain atrophy that were represented across the diagnostic groups. Canonical correlation analysis of clinical and imaging components found three key brain-behaviour relationships that revealed a continuous spectrum across the cohort rather than discrete diagnostic entities. In the forty-six patients with longitudinal follow up (mean 3.6 years) syndromic overlap increased with time. Together, these results show that syndromes associated with FTLD do not form discrete mutually exclusive categories from their clinical features or structural brain changes, but instead exist in a multidimensional spectrum. Patients often manifest diagnostic features of multiple disorders and deficits in behaviour, movement and language domains are not confined to specific diagnostic groups. It is important to recognise individual differences in clinical phenotype, both for clinical management and to understand pathogenic mechanisms. We suggest that the adoption of a transdiagnostic approach to the spectrum of FTLD syndromes provides a useful framework with which to understand disease progression, heterogeneity and treatment.

## Introduction

The clinical disorders caused by frontotemporal lobar degeneration pathologies (FTLD) are highly heterogeneous in their pathology and phenotypes (Kertesz *et al*., 2005; MacKenzie *et al*., 2010; Rohrer *et al*., 2011). Patients are typically diagnosed as having one of several principal syndromes, including behavioural variant frontotemporal dementia (bvFTD)(Rascovsky *et al*., 2011), primary progressive aphasia (with the non-fluent nfvPPA and semantic svPPA subtypes)(Gorno-Tempini *et al*., 2011), progressive supranuclear palsy (PSP)(Höglinger *et al*., 2017) or corticobasal syndrome (CBS)(Armstrong *et al*., 2013). The clinicopathological correlations of these syndromes are imprecise (Irwin *et al*., 2015). For example, bvFTD can be associated with Tau, TDP-43, or FUS protein inclusions or mixed neuropathology (Perry *et al*., 2017). Some clinical syndromes, such as PSP-Richardson’s Syndrome, have good correlation with the associated pathology (Gazzina *et al*., 2019), however the corresponding pathology may have diverse phenotypic expressions (Respondek *et al*., 2014). Recent revisions of diagnostic criteria recognise this heterogeneity (Armstrong *et al*., 2013; Höglinger *et al*., 2017), and there may be future improvements in clinicopathological correlations by imaging or fluid-based biomarkers, aiming to optimise patient selection for disease modifying therapies (Irwin *et al*., 2015; Meeter *et al*., 2017).

Here we propose that the effort to refine diagnostic segregation of the disorders has fundamental limitations. These are not merely due to the limits of a given test or biomarker but are biologically real constraints that can in turn be informative about the nature of the disorders. We suggest that a better understanding of aetiology and pathophysiology, and more effective therapies, can be gained by examining the phenotypic patterns across the broad spectrum of all FTLD-associated disease. Symptomatic therapies may especially benefit from such a transdiagnostic approach, selecting patients based on the presence of relevant clinical features, whichever their diagnostic label or proteinopathy.

A transdiagnostic approach is increasingly used in psychiatry, epitomised by the Research Domain Criteria methodology (Kozak and Cuthbert, 2016; Grisanzio *et al*., 2018). A similar approach is applicable to neurodegenerative diseases with overlapping phenotypes (Lambon Ralph *et al*., 2003; Husain, 2017) and cognitive deficits after stroke (Butler *et al*., 2014; Mirman *et al*., 2015; Halai *et al*., 2017). There are many overlapping symptoms and indistinct phenotypic boundaries between FTLD syndromes (Kertesz *et al*., 1999, 2005). For example, executive dysfunction is a common cognitive impairment across FTLD-associated syndromes (Burrell *et al*., 2014; K G Ranasinghe *et al*., 2016) and changes in behaviour, social cognition and personality, while characteristic of bvFTD are also seen in PSP (Cordato *et al*., 2005; Ghosh *et al*., 2012; Gerstenecker *et al*., 2013), CBS (Huey *et al*., 2009) and the primary progressive aphasias (Rosen *et al*., 2006; Rohrer and Warren, 2010). Neuropsychiatric symptoms, including apathy and impulsivity, occur in multiple FTLD syndromes (J. Rohrer *et al*., 2010; Lansdall *et al*., 2017). The movement disorders typical of PSP and CBS can also develop in patients diagnosed with bvFTD (Park *et al*., 2017) and nfvPPA (Santos-Santos *et al*., 2016). Language impairments are seen across all FTLD syndromes, including bvFTD (Hardy *et al*., 2015), PSP and CBS (Peterson *et al*., 2019).

We therefore used a transdiagnostic approach to assess the phenotype of FTLD syndromes. We tested the hypothesis that syndromes associated with FTLD are multidimensional clinical spectra, rather than discrete clinical entities. A colour-map symbolises the current most widely used approach, in which patients have a distinct clinical phenotype of a singular syndrome, represented by a discrete colour patch (Figure 1A: ‘red bvFTD’ is distinct from ‘blue PSP’) (Butler *et al*., 2014). Our alternate hypothesis is that patients lie in a continuous colour-space, shown in in Figure 1B. Intermediate or mixed phenotypes like PSP-F, CBS-NAV or svPPA with prominent behavioural disturbance, are readily placed within the continuous phenotypic space. A corollary hypothesis is that the multivariate clinical spectrum of the disorders can be mapped to multivariate regional structural brain change. Note that this is not an argument for ‘lumping’ patients into super-ordinate diagnostic groups, or for ‘splitting’ diagnoses into ever finer subtypes. This type of transdiagnostic approach recognises the clear individual differences across patients and does not propose an unstructured pool; instead, the key hypothesis is that the underlying variations in FLTD reflect a statistical structure in the form of multiple graded dimensions rather than mutually exclusive categories. Thus, the concept of phenotypic spectra allows for both the recognition of broad similarities and unique combinations of features.

**Figure 1.**
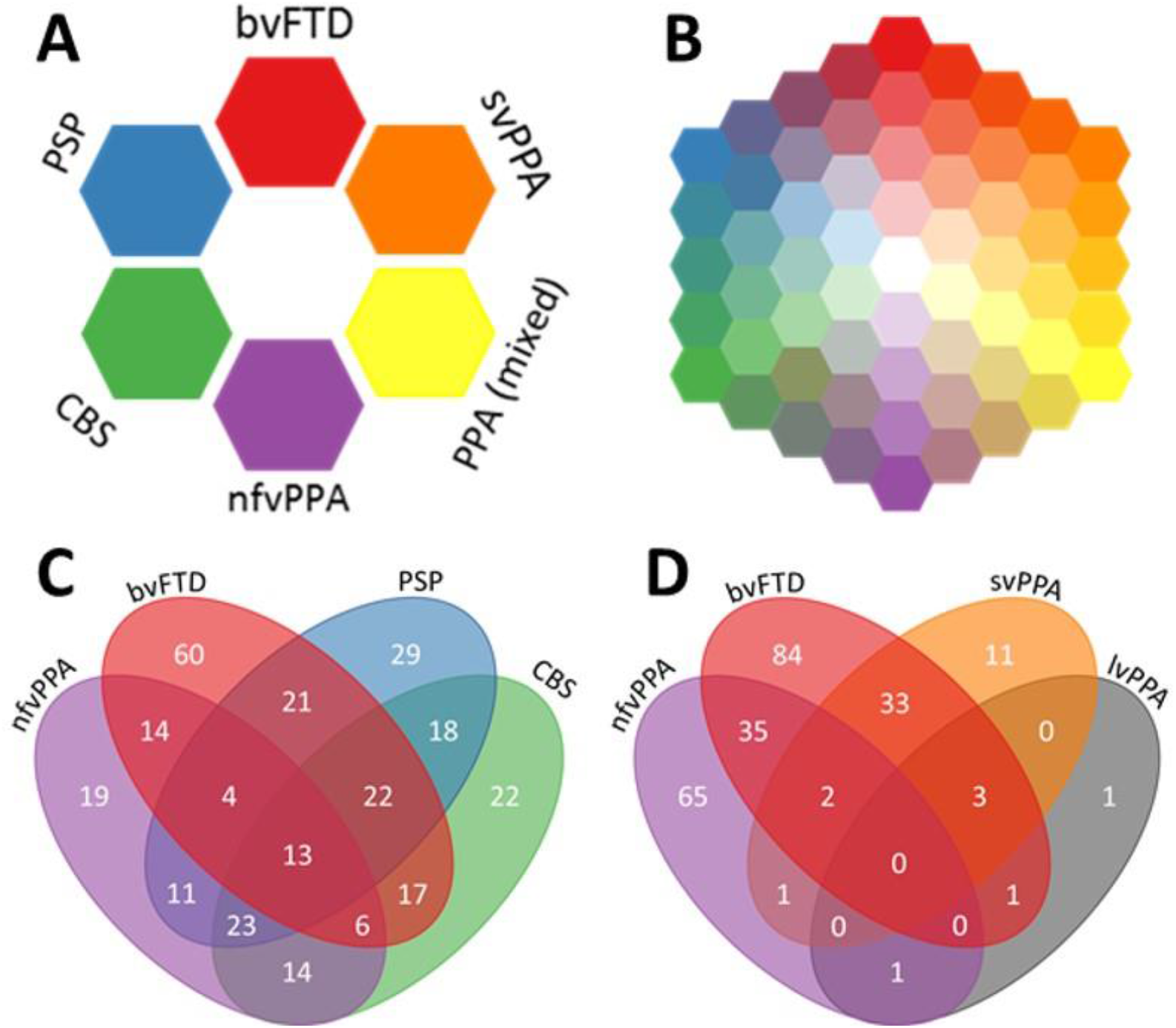
The FTLD syndrome spectrum. 1A: Schematic of current diagnostic criteria. 1B: Schematic to highlight our hypothesis that FTLD syndromes occur on a spectrum. 1C and D: Four-way Venn diagrams of overlap between FTLD syndromes in the study. The numbers in each oval refer to the number of patients who met the diagnostic criteria for those syndromes. Many patients met the diagnostic criteria for two or more syndromes. 1C: Overlap between bvFTD, nfvPPA, PSP and CBS. 1D: Overlap between bvFTD, nfvPPA, svPPA and lvPPA.

To test our hypotheses, we exploited the epidemiologically-based PiPPIN dataset (Coyle-Gilchrist *et al*., 2016) and a replication dataset acquired four years later. We undertook a systematic behavioural, cognitive and imaging assessment of patients with syndromes associated with FTLD, in a region of 1.75 million people in the United Kingdom. We predicted that while classical syndromes of bvFTD, PPA, PSP and CBS exist, a data-driven approach would reveal phenotypic continuity without clear separation between phenotypes. With longitudinal follow up of a subset of participants, we tested the hypothesis that clinical phenotypes merge by addition of features, with increasing overlap – analogous to the move towards the centre of the colour-space. Moreover, we predicted that clusters of symptoms would be associated with a specific pattern of brain atrophy, while the extent to which a patient has this atrophy pattern determines the severity of the associated symptoms.

## Materials and methods

The rapidly evolving field of FTLD/FTD/PSP research can result in confusion in definitions and diagnostic labels. In this paper we use the current consensus nosology for clinical and pathological diagnoses. We use frontotemporal lobar degeneration (FTLD) to refer to the pathology, subtyping to tau or TDP43 pathologies where applicable. The phrase “Frontotemporal lobar degeneration syndromes” refers collectively to the clinical diagnoses of bvFTD (with or without motor neuron disease), PPA, nfvPPA, svPPA, PSP or CBS and their intermediate phenotypes. The term corticobasal degeneration is limited to the pathology, while corticobasal syndrome (CBS) refers to the clinical syndrome. Note that not all patients will have FTLD pathology (especially lvPPA and mixed PPA patients) and not all those with FTLD pathology will have had one of the corresponding syndromes.

### Participant recruitment

The “Pick’s Disease and Progressive Supranuclear Palsy Prevalence and Incidence study” (PiPPIN) sought to recruit all patients with a clinical diagnosis of a frontotemporal lobar degeneration syndrome living in the counties of Cambridgeshire and Norfolk in the United Kingdom. Cross sectional assessments were performed during two 24-month periods, from 1^st^ January 2013 to 31^st^ December 2014 and again from 1^st^ January 2017 to 31^st^ December 2018. Participants were recruited via multiple routes, including specialist cognitive and movement disorder clinics at tertiary and secondary healthcare services (using paper and electronic health records), patient support groups (FTD support group, PSP Association), advertisements in local newspapers and through local research databases and the National Institute for Health Research “Join Dementia Research” registry. Patients were recruited at all disease stages. We sought to assess all participants, either at our research centre or at their home or care home. Patients alive during both study periods were invited to assessment in both periods, but only their first visit was used for the cross-sectional analysis. 365 patients were identified in the catchment area, 310 of whom were met in person by the study team for phenotypic assessment. Death or end-stage disease were the main reasons for our not assessing the remaining 55 cases. All participants provided written informed consent or, if they lacked capacity to consent, their next of kin was consulted using the “personal consultee” process established by UK law. The study had ethical approval from the Cambridge Central Research Ethics Committee (REC 12/EE/0475).

### Clinical assessment

We used a structured clinical assessment to record the presence or absence of symptoms and signs typically seen in frontotemporal lobar degeneration syndromes. This assessment contained all clinical features included in the current consensus diagnostic criteria (Supplementary Information) (Rascovsky *et al*., 2007; Bensimon *et al*., 2009; Gorno-Tempini *et al*., 2011; Armstrong *et al*., 2013; Höglinger *et al*., 2017). Each patient’s primary diagnosis was made according to these criteria, with reference to the dominant features at the time of presentation and assessment. Patients with a mixed primary progressive aphasia, who met the diagnostic criteria for PPA but not one of the three subtypes (Gorno-Tempini *et al*., 2011) were grouped with lvPPA for this study, in view of the low numbers and the association of both phenotypes with Alzheimer’s pathology (Sajjadi *et al*., 2012). For patients who met several sub-diagnostic criteria we grouped disease probable and possible diagnoses together, and classified by the dominant phenotype and formal MAX-rules where available (Grimm *et al*., 2019). We re-applied the other diagnostic criteria to each patient to assess if he or she met the diagnostic criteria for any of the other FTLD syndromes (excepting the ‘mutual exclusivity’ clause included in several criteria). Patients completed the Addenbrooke’s Cognitive Examination wherever possible (ACER) (Mioshi *et al*., 2006) and a carer’s assessment was obtained using the Cambridge Behavioural Inventory (CBI-R) (Wear *et al*., 2008). At the time of writing, forty-nine participants have undergone *post mortem* examination, via the Cambridge Brain Bank.

### Imaging analysis

One hundred and thirty-three patients (bvFTD n=28, nfvPPA n=15, svPPA n=5 PPA n=10, PSP n=53, CBS n=22) from the phenotyped cohort were scanned at the Wolfson Brain Imaging Centre, University of Cambridge on a Siemens 3T system. Structural magnetic resonance imaging was performed using a T_1_-weighted magnetisation-prepared rapid acquisition gradient echo (MPRAGE) sequence. Images were pre-processed using SPM12 with default settings. Grey and white matter segments were combined to whole brain images for further analysis. The DARTEL pipeline was used to create a study specific template using all images. Age and total intracranial volume (TIV) were included in a multiple regression and regressed out of the data. Source based morphometry was used on the residual images to identify covarying networks of grey and white matter atrophy, further details of this step are given in the next paragraph.

### Statistical analysis

A summary of the analysis pipeline is shown in Figure 2. First, we examined the relationships between individual clinical features using distance measures and multidimensional scaling (Shepard, 1980) (Figure 3). The pairwise Jaccard’s distances between clinical features were calculated, resulting in a dissimilarity matrix. Non-classical two-dimensional scaling was performed on this dissimilarity matrix (Shepard, 1980).

**Figure 2.**
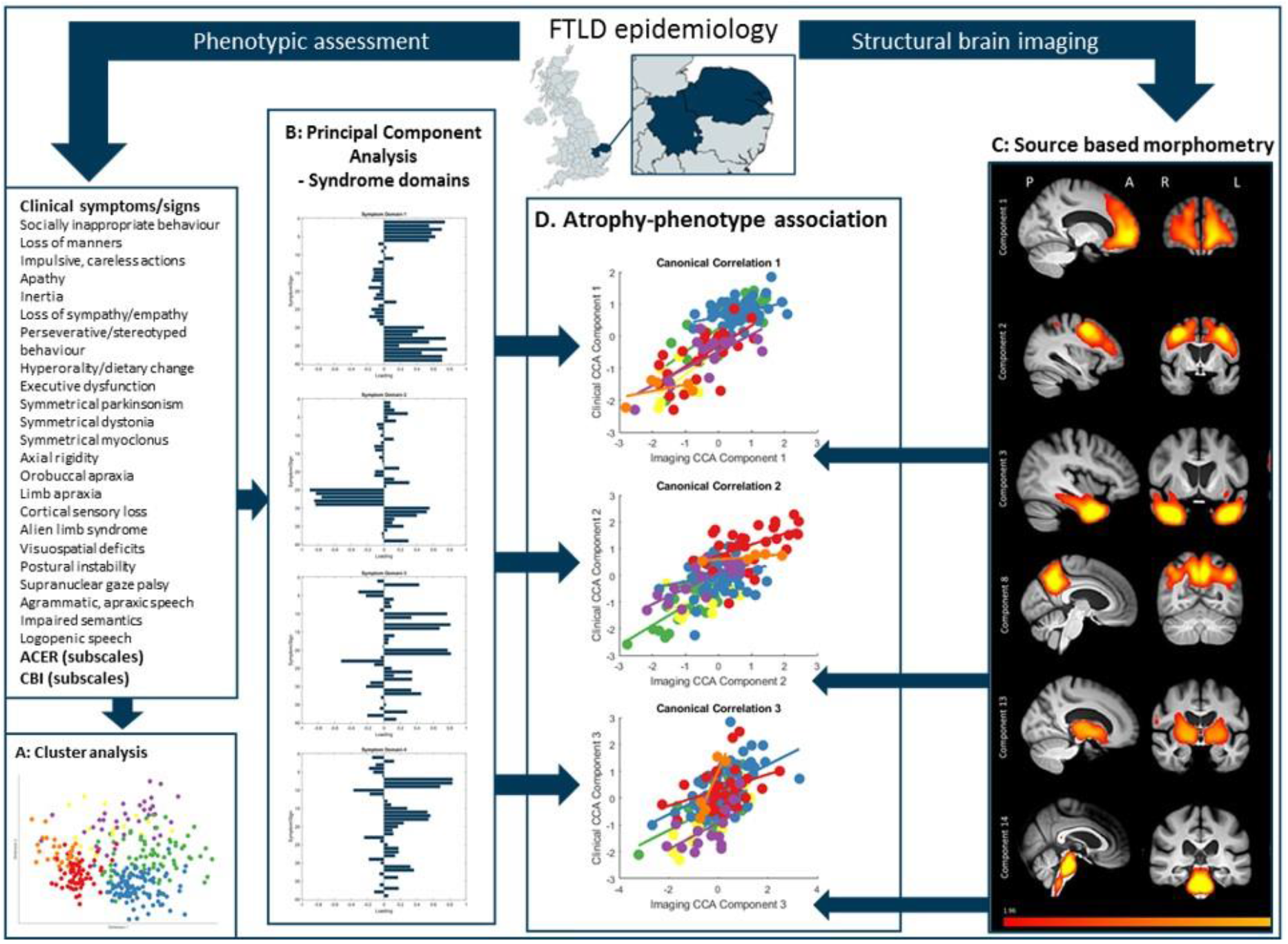
Schematic of data processing. First, patients were recruited from the study catchment area for phenotypic assessment and structural brain imaging. Second, a cluster analysis was performed on clinical features. Third, we performed a principal component analysis on all clinical features to find latent syndrome dimensions across FTLD. Fourth, we used source based morphometry (independent component analysis on grey and white matter) created atrophy components. Finally, we then explored the relationship between phenotype (syndrome dimensions from the principal component analysis) and brain structure (source-based morphometry imaging components) using canonical correlation analysis.

**Figure 3.**
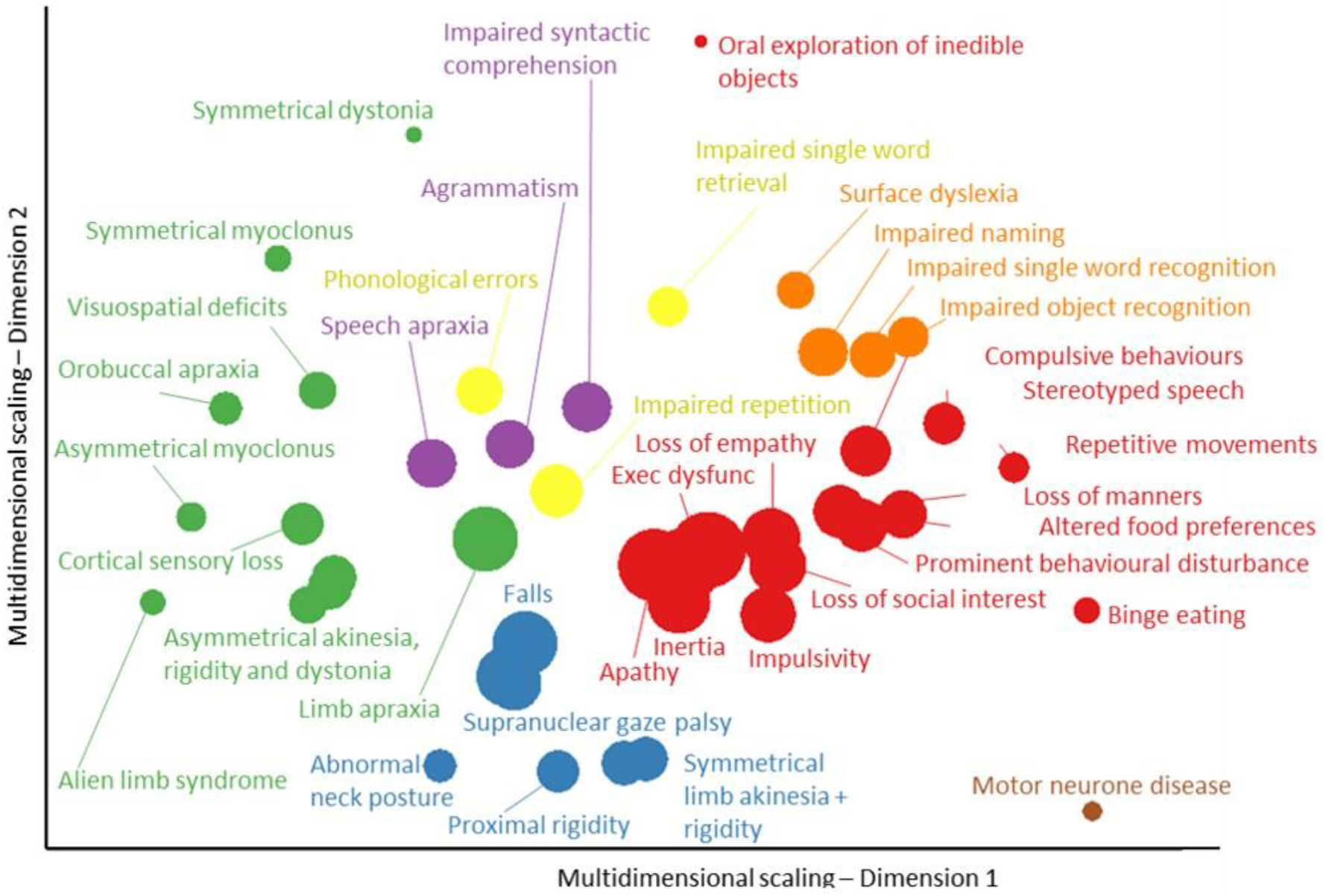
Cluster analysis and multidimensional scaling of behavioural, language and motor impairments in FTLD. Each feature is colour coded by FTLD subtype (same colour codes as Figure 1) based on the primary diagnostic criteria which the symptom contributes to. The size of each point is scaled based on its prevalence in the cohort (larger icons have a higher prevalence). Symptoms from each FTLD syndrome cluster together, but many features are also closely located to those from other syndromes.

**Figure 4.**
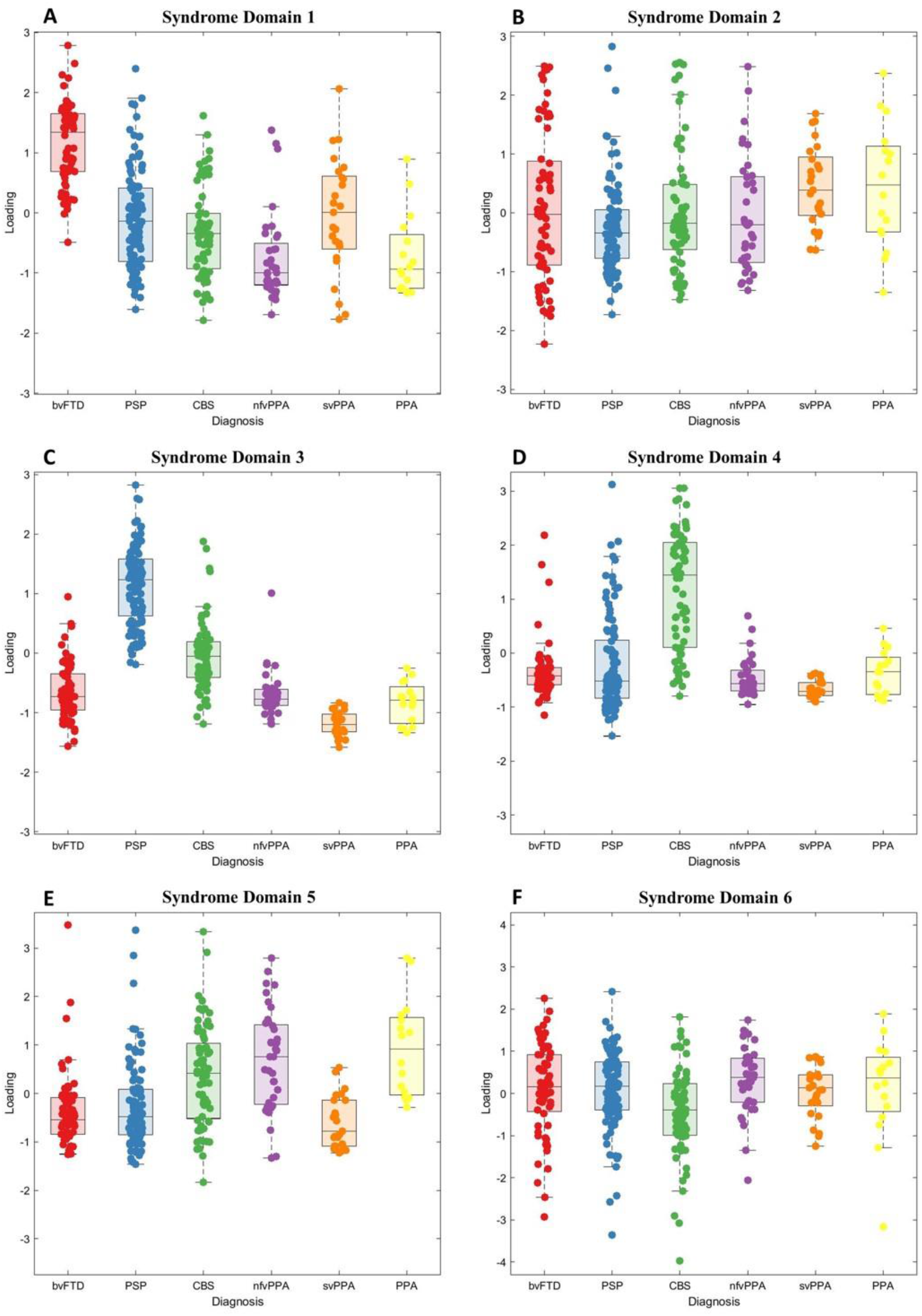
Principal component analysis scores of clinical features in FTLD syndromes. Six principal components (4A-F) were selected. 4A: Syndrome dimension 1 - clinician and carer ratings of behavioural impairment. 4B: Syndrome dimension 2 - global cognitive impairment, comprised of all Addenbrooke’s Cognitive Examination subscores. 4C: Syndrome dimension 3 - supranuclear gaze palsy, postural stability and symmetrical rigidity (positive loading) and semantic language impairment (negative loading). 4D: Syndrome dimension 4 - asymmetrical parkinsonism, dystonia, myoclonus with limb apraxia, cortical sensory loss and alien limb syndrome. 4E: Syndrome dimension 5 - agrammatic, apraxic and logopenic language impairments. 4F: Syndrome dimension 6 - carer ratings of low mood and abnormal beliefs.

Second, we looked for patterns of covariation in the presence or absence of clinical features (Perry *et al*., 2017; Grisanzio *et al*., 2018; Schumacher *et al*., 2019). We grouped the forty-five binary clinical features into twenty-four groups of related features by summing the number of present features in each group. Clinical feature groups were defined by categories in the diagnostic criteria. A full list of clinical symptoms and signs and their groupings is shown in supplementary materials. The clinical feature group scores, ACER and CBIR results were standardised into z scores then entered into a principal component analysis. A Kaiser-Meyer-Olkin test was used to determine the suitability of our dataset for principal component analysis. We selected six components using Cattell’s criteria then performed varimax rotation.

Third, we characterised patterns of covariation in grey and white matter atrophy across all participants. We used the GIFT software package to perform source based morphometry, a multivariate alternative to voxel based morphometry which uses independent component analysis (Xu *et al*., 2009). Source based morphometry was performed on the residual values from the pre-processed images (see imaging analysis section of methods for details). We extracted 15 independent components of covarying brain atrophy (Figure 5), and confirmed the reliability of these components using ICASSO with 100 repetitions (Himberg and Hyvärinen, 2003).

**Figure 5.**
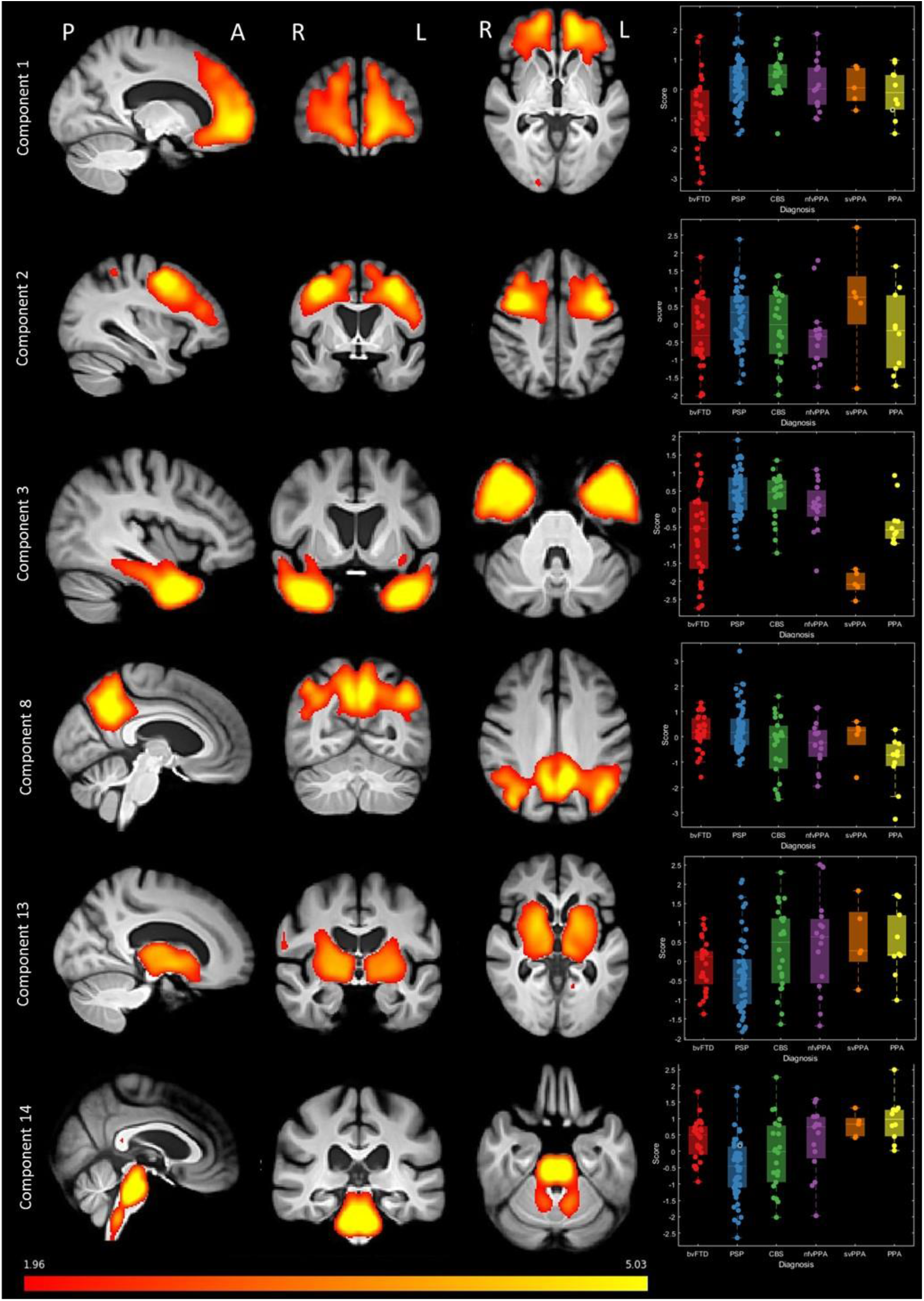
Source based morphometry (based on independent component analysis) of combined grey and white matter. A subset of components is shown here, all components are shown in supplementary materials. A total of 15 components were selected, each representing a region of independently covarying grey and white matter atrophy. Images are standardised group spatial maps for each component, superimposed on an average of all brain images. The scatter-box plots show the standardised subject loading coefficients, grouped by FTLD syndrome subtype.

Fourth, we looked at the relationship between clinical phenotype and brain atrophy (Figure 6). We used canonical correlation analysis (CCA) to identify canonical variates between the six principal components from the clinical feature data (Figure 4) and the fifteen components from the imaging analysis (Figure 5) (Tsvetanov *et al*., 2018). All inputs were standardised into z scores before CCA. Pearson’s correlations were corrected for multiple comparisons by estimating the false discovery rate using the *mafdr* function in Matlab 2018b.

**Figure 6.**
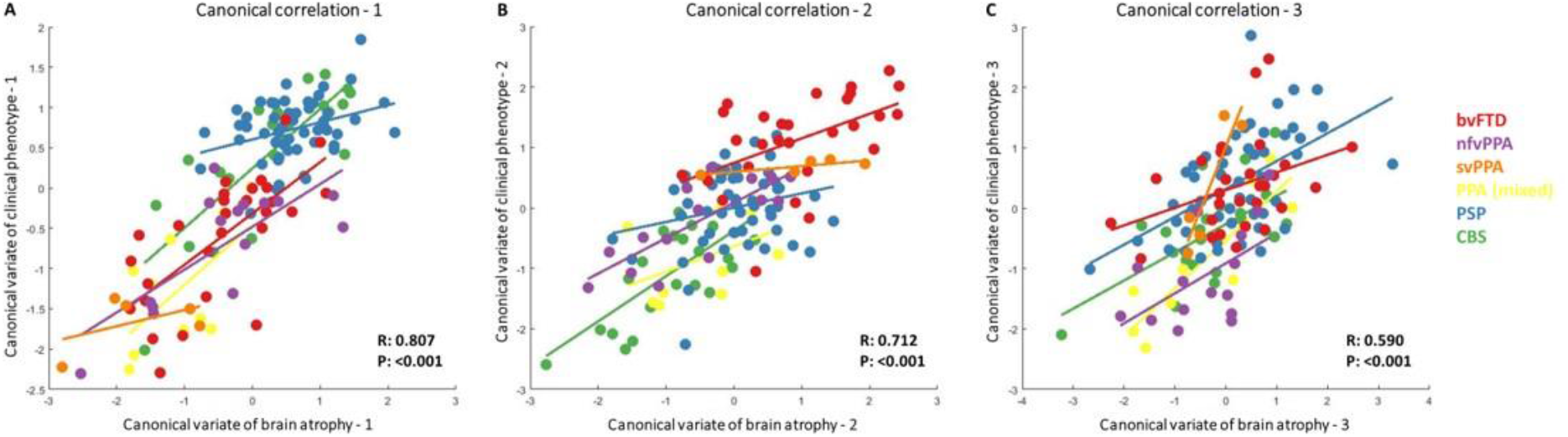
Structure-phenotype associations using canonical correlation analysis with phenotypic (syndrome dimensions from principal component analysis) and structural (atrophy components from source based morphometry) information. Three canonical correlation components were selected, each comprised of multiple imaging and clinical phenotype components. 6A: First canonical correlation. Atrophy in the motor cortex and brainstem had the greatest loading onto the imaging component. Syndrome dimensions 3 (PSP-like motor features) and 4 (CBS-like motor features) had positive loadings and syndrome dimension 2 (global cognitive impairment) had negative loading on the clinical component. 6B: Second canonical correlation. Atrophy in the frontal and temporal lobes had the greatest loading on the imaging component. On the clinical component, syndrome dimension one (behavioural impairment) had positive loadings. 6C: Third canonical correlation. A spread of cortical and subcortical atrophy components loaded on the imaging component and syndrome dimensions 1, 2 and 3 contributed to the clinical component. Plots of loadings onto all imaging and clinical components is shown in Supplementary Materials.

Finally, we looked at longitudinal change in clinical feature component scores in the subset of patients (n=46) who were reviewed twice. We converted follow up participant scores into zscores based on the baseline data, by matching each score to the respective z-score in the baseline data. This ensured follow up participants were matched to the cross-sectional dataset. We then multiplied these standardised follow-up z scores by the baseline principal component coefficients to get follow-up principal component scores.

All patients had a clinical phenotypic assessment but other measures (including ACER and CBI-R) were subject to missing data. Missing data (6.32% of the total dataset) were imputed using trimmed scored regression (Folch-Fortuny *et al*., 2016) using the partial dataset of that participant as predictors. All statistical and imaging analysis was performed in Matlab 2018b (Mathworks, USA) apart from ANOVA and Chi squared tests which were performed in JASP (version 0.9.2).

### Data Availability

Anonymised data are available on reasonable request for academic purposes.

## Results

A detailed epidemiological assessment of FTLD syndromes in the study area has previously been reported (Coyle-Gilchrist *et al*., 2016). Further demographic details of the study cohort, including the later recruitment period, are shown in Table 1.

**Table 1.**
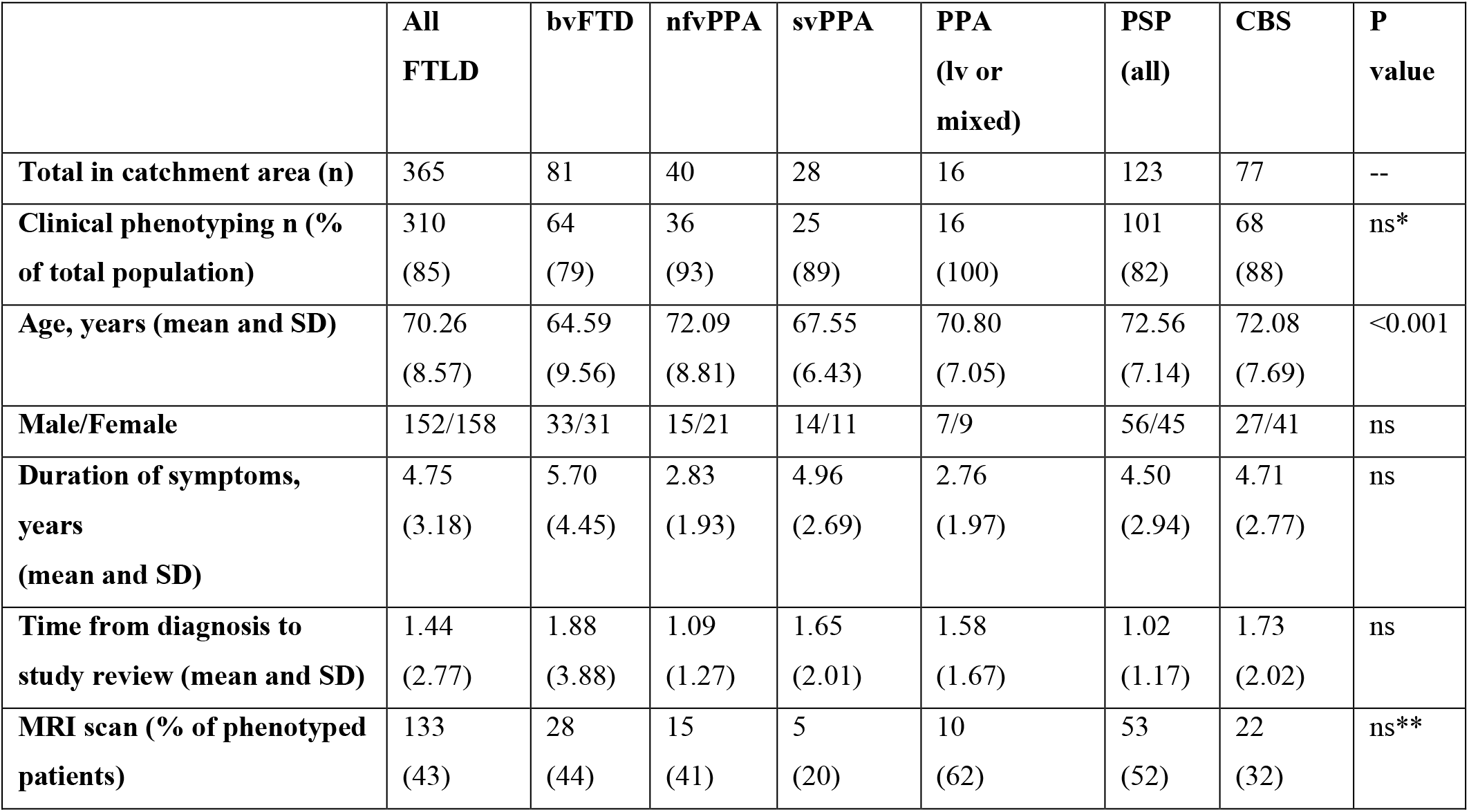
Demographics of the study cohort. P values are the result of ANOVA or Chi squared test for each row on FTLD subgroups, ns= not significant (p>0.05), *ANOVA of percentage of total population in each group, ** ANOVA of percentage of phenotyped patients in each group.

We assessed in person 85% (310/365) of the patients identified as living in the study catchment area with a FTLD syndrome. Fifty eight patients had a definite diagnosis of FTLD, either by subsequent *post mortem* pathological diagnosis (n=49) or a causative genetic mutation on clinical genetics tests. Of patients who underwent *post mortem* examination, all patients with a clinical diagnosis of PSP had PSP pathology (n=14). Most patients with svPPA had FTLD-TDP-43C (n=3/4), one had Pick’s pathology. bvFTD was associated with FTLD-tau (Picks’ n=1, PSP n=1) or TDP43 (n=5f). The majority of patients with CBS had either CBD (n=6/19) or Alzheimer’s disease (8/19). Of the four patients with nfvPPA three had FTLD-tau and one had Alzheimer’s disease. The one patient with lvPPA had Alzheimer’s Disease pathology.

Sixty-two percent of patients (n=194) met the core diagnostic criteria for more than one syndrome, with patients meeting the inclusion criteria for two (n=112), three (n=69) or four (n=13) diagnoses (Figure 1C and D). The most commonly overlapping syndromes were PSP and CBS (n=76), bvFTD and either PSP (n=60) or svPPA (n=38) and nfvPPA with either CBS (n=56) or PSP (n=51).

We used cluster analysis to investigate how closely clinical features related to each other. Multidimensional scaling of clinical features (across all patients) broadly recapitulated the phenotypic clustering as represented by the classical phenotypes of each syndrome (Figure 3). However, there were also many close links between signs conventionally associated with distinct diagnoses. For example, progressive behavioural change, apathy, inertia and impulsivity (typical of bvFTD), were close to symmetrical parkinsonism, falls, axial rigidity and a supranuclear gaze palsy (typical of PSP). Other features suggestive of bvFTD (socially inappropriate and compulsive behaviour and stereotypy of speech), were close to features typical of svPPA features (impaired naming, single word comprehension and object recognition). PSP and CBS features were closely linked, while speech apraxia, agrammatism and impaired syntactic comprehension (indicative of nfvPPA) overlapped with limb apraxia (indicative of CBS).

First, we sought latent syndromic dimensions using principal component analysis of the phenotypic data. Six principal components were identified using Cattell’s criteria, each representing a group of covarying features (encompassing symptoms, signs, ACER and CBI scores, varimax-rotated component matrix in Supplementary Materials). These six components explained 58.52% of the variance in the dataset (Kaiser-Meyer-Olkin=0.86). Syndrome dimension 1 (Figure 4A) reflected clinician and carer ratings of behaviour and personality change, with executive dysfunction, impulsivity and disinhibition, loss of empathy, stereotyped behaviours, hyperorality and dietary change, apathy, endorsements of abnormal behaviour, altered eating habits and stereotypic and motor behaviour subscales. This “behaviour” dimension was expressed strongly by patients with bvFTD, but also a high proportion of PSP, CBS and svPPA patients. Some patients in these latter groups had weightings similar to bvFTD. The second syndrome dimension (Figure 4B) reflected global cognitive function, with negative loadings from ACER subscores. Carer ratings of everyday function and memory also had positive loading onto this dimension (higher CBI score, reflecting greater impairment). There was wide variation in this dimension’s weighting across all groups, with higher scores reflecting worse cognitive impairment.

The third dimension (Figure 4C) reflected axial rigidity, postural instability and a supranuclear gaze palsy (positive loading) in the absence semantic language impairments (negative loading). Thus, patients with typical PSP and typical svPPA lie at opposite ends of this dimension, with high and low scores respectively. However other groups had a spread of scores, many patients with corticobasal syndrome had very high scores (PSP-like). Some bvFTD had high scores indicating a PSP-overlap, while others had low scores, implying presence of semantic impairment.

Positive scores on syndrome dimension four (Figure 4D) represented asymmetrical parkinsonism, dystonia and myoclonus with cortical features of apraxia, cortical sensory loss and alien limb syndrome. Patients with corticobasal syndrome, and a subset of patients with PSP had high scores in this dimension. Dimension 5 (Figure 4E) represented language impairments, agrammatic, apraxic and logopenic speech with motor features (myoclonus and limb apraxia). Patients with CBS, nfvPPA, logopenic variant and mixed PPA had high weighting on this dimension, as did a small subset of those with clinical diagnoses of PSP and bvFTD. Dimension 6 explained less variance than the other components and represented primarily carer ratings of mood and abnormal beliefs (Figure 4F).

Second, we investigated the structural changes associated with FTLD, and their associations with the clinically orientated syndromic dimensions. The scanned subset of participants was similar to the whole population, with no statistically significant differences in weighting for dimensions 1,2,3,5, and 6. The differences in syndrome dimension 2 (t=2.41, p=0.016) indicated less severe global cognitive impairment in those who were scanned. Source based morphometry revealed fifteen significant structural components, each representing a pattern of covaried atrophy (subset in Figure 5, all components in Supplementary Materials). The components had high stability across 100 ICASSO runs (mean=0.981, standard deviation=0.004). The loadings on these imaging components were not confined to single diagnostic groups.

Imaging components one and two related to the frontal and prefrontal cortex; patients with bvFTD tended to have low scores on these components (i.e. atrophy), but many patients with nfvPPA, PSP and CBS also had low scores indicating a frontal cortical atrophy (Figure 5). Component three, with bitemporal atrophy, had very strong negative scores in all svPPA patients, but also many bvFTD patients. Some participants with CBS, nfvPPA and PPA had negative scores on imaging component eight, which reflected biparietal atrophy. Imaging component thirteen represented the volumes of corticospinal tracts and basal ganglia. Many patients with PSP, but also some patients with bvFTD, CBS and nfvPPA had low scores on this component. Component fourteen represented brainstem atrophy, with large negative scores in PSP and CBS but also some nfvPPA patients.

Third, we looked for structure-function correlations between the clinical and imaging components. Since both cognition and atrophy are intrinsically multivariate, we used canonical correlation analysis between the six cognitive dimension and fifteen atrophy components. Three canonical correlations were selected for further analysis (each p<0.05, rejecting the null hypothesis that the canonical correlation is zero). The first canonical correlation (R=0.81, p<0.001) represented the association between motor impairments (syndrome dimensions three and four) and relatively preserved cognition (syndrome dimension two) with motor cortex and brain stem atrophy (atrophy components six and fourteen). Patients with PSP, CBS and some patients with bvFTD had positive loadings, while patients with primary progressive aphasia (notably the svPPA subtype) and some with bvFTD had negative loadings (Figure 6A). Four of the six FTLD subgroups had significant correlations in this canonical correlation: PSP (Pearson’s R:0.33, p:0.03), CBS (R:0.81 p:<0.001), bvFTD R:0.70 p:<0.001) and nfvPPA (R:0.74 p:0.03) (All results in Supplementary Materials).

The second canonical correlation (R=0.71, p<0.001) represented the spectrum between behavioural impairment (syndrome dimension 1) associated with atrophy in the frontal and temporal lobes (atrophy components 1 and 3) versus global cognitive impairment, apraxia, cortical sensory loss and language impairments in association with atrophy in the parietal cortex (atrophy components 7 and 8). Positive weightings on this canonical correlation were most common for bvFTD, svPPA and a subset of PSP. Negative loadings were predominantly seen in CBS and a few patients with nfvPPA and mixed PPA (Figure 6B). bvFTD (R=0.49, p=0.02), nfvPPA (R=0.79 p=0.001) and CBS (R=0.7, p=0.001) most contributed to this canonical variate.

The third canonical correlation (R=0.58 p<0.001) represented a combination of behavioural, cognitive and motor symptoms in association with atrophy in motor and parietal cortices, basal ganglia and brainstem (Figure 6C). This canonical correlation had positive loadings across a wide range of diagnoses, with no clear group separation. This canonical variate was driven by CBS (R=0.62 p=0.005), PSP (R=0.54 p<0.001) and PPA (R=0.87 p=0.002) subgroups with a weaker contribution from svPPA (R=0.91, p=0.048). The three residual, unselected canonical covariates did not correlate in any FTLD subgroup.

The final analysis considered the longitudinal change in the forty-six patients who were alive and assessed in both 2013-2014 and 2017-2018. The mean time between assessments was 3.6 years (standard deviation 0.87 years). Between first and second assessments there was progression in all syndrome dimensions across all groups. At the second assessment there was greater overlap between diagnostic groups, across all syndrome dimensions (Figure 7). More patients met two or more sets of diagnostic criteria (after removing mutual exclusivity criterion) at follow up (n=42) compared to baseline (n=33) (Chi squared statistic with Yates correction 4.618, p=0.031).

**Figure 7.**
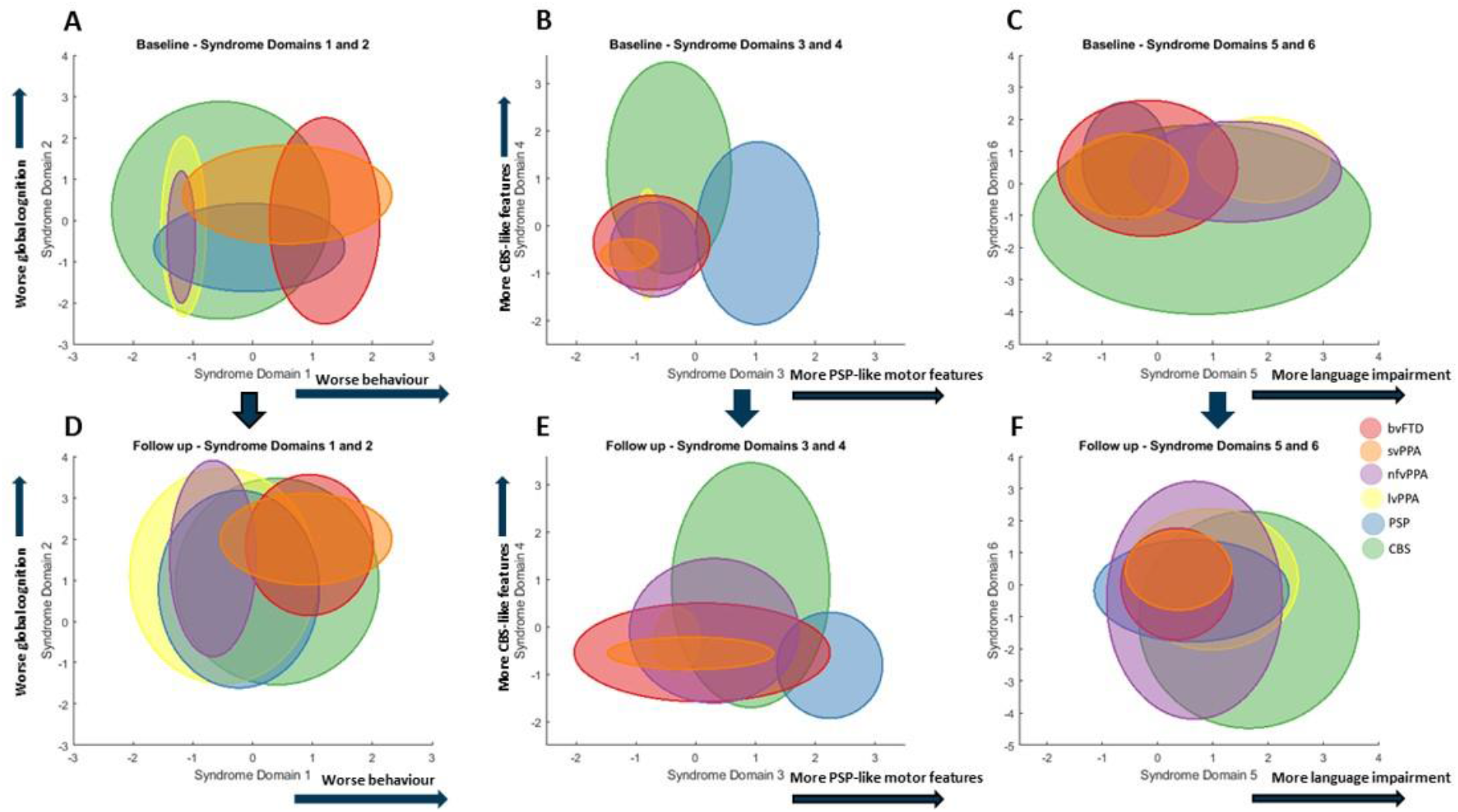
Longitudinal phenotype information. A subset of patients were assessed at two timepoints. Each circle shows the 95% confidence intervals of the syndrome dimension scores for each FTLD subgroup at baseline and follow up. At follow up there was greater overlap across all FTLD syndromes in all syndrome dimensions

## Discussion

This epidemiologically based study used a data-driven analysis of cross-sectional phenotypes to show that the common syndromes associated with frontotemporal lobar degeneration are not discrete in their clinical features or structural brain changes (Figure 1A), but instead exist as a multidimensional spectrum (Figure 1B). Many patients displayed the diagnostic features for multiple diagnoses (Figure 1C&D). The dimensions of behaviour, movement and language features occur to varying degrees across all the major diagnostic groups. Differences between groups were expressed by different weightings along these spectra, rather than by categorical clinical or imaging features.

Despite the continuity among patient phenotypes, the clinical syndromes are not random associations. Our analyses revealed close associations between sets of cognitive, behavioural, language and motor symptoms and signs which are reminiscent of the classical phenotypes (Figure 2). For example, syndrome dimension three, which represents a supranuclear gaze palsy, falls, akinesia and preserved semantics, is readily identified as a pattern typical of PSP-Richardson’s syndrome. However, forty-four percent of CBS patients expressed this pattern to the same degree as PSP patients. The recognition of such overlap has contributed to the development of intermediate diagnoses like PSP-CBS (Höglinger *et al*., 2017) and CBS-PSP (Armstrong *et al*., 2013) but our results indicate that such overlap is common rather than exceptional. However not all potential intermediate phenotypes occur. For example, a supranuclear gaze palsy, axial and symmetrical limb rigidity rarely coexist with semantic impairment, a combination which has only rarely been reported in patients with mixed tau and TDP43 pathology (Snowden *et al*., 2019).

We propose that this spectral approach is critical to understand the biological basis of the complex clinical syndromes, and to target future therapies appropriately. Rather than focus on the determinants of disease or its treatment by diagnosis, one can focus on the determinants and treatment of the syndromic dimensions, in whichever diagnostic ‘group’ these dimensions are expressed. To do otherwise risks the misdirection of a treatment or the dilution of the effects of aetiological factors, whether genetic, environmental, or aggregate of pathogenic proteins. In other words, one could understand and potentially treat the “PSP-like” features whether they occurred in the context of clinically diagnosed PSP-Richardson’s syndrome, CBS or bvFTD.

We do not suggest that the current diagnostic criteria are invalid. Instead, our results highlight the limitations of a categorical approach to diagnosis when the disorders are inherently multivariate spectra in their clinical and imaging features. Nor do our data suggest incorrect diagnosis: although only forty nine patients in this study have had post mortem examination, they confirmed clinicopathological correlations in keeping with the literature (very high for PSP (Gazzina *et al*., 2019) and svPPA (Spinelli *et al*., 2017), predominantly CBD or AD pathologies for CBS (Alexander *et al*., 2014), and either Tau or TDP43 pathologies for bvFTD (Perry *et al*., 2017)). Indeed, the symptom-based data-driven cluster analysis broadly reproduced the diagnostic criteria. But, the relative weightings on such clusters were graded, which highlights the difficulties when applying diagnostic criteria to some patients, especially those with intermediate or mixed phenotypes.

In our analysis, we did not differentiate features that are more salient to a clinician (e.g. supranuclear gaze palsy) from those that are more salient to a relative or carer (e.g. behavioural disturbance, non-fluent aphasia or falls). This difference in perspective is relevant to diagnostic labelling. For example, a patient with apraxia, akinesia, dystonia and non-fluent agrammatic speech might be diagnosed as CBS or nfvPPA according to the dominant clinical features, but whose opinion on dominance matters most, the patient, carer or clinician? This is complicated further by the change in insight associated with many FTLD syndromes (O’Keeffe *et al*., 2007). A further complication for the categorical approach to diagnosis is the evolution of behavioural, motor or language features over time which raises the question of whether the diagnosis label should be changed or complimented by a secondary, parallel diagnosis. Our approach largely resolves this issue by taking a transdiagnostic approach based on clinical and/or imaging domains, which we consider in the next section.

Our data-driven approach identified close clustering of the clinical features and six latent syndrome dimensions that demonstrated the high degree of overlap across FTLD syndromes. Behavioural features were closely clustered and loaded onto one syndrome dimension. However, they also clustered near cognitive and motor symptoms/signs. Apathy and impulsivity had a close link, reflecting the fact that they often coexist, rather than representing opposite ends of a hyper-hypo-kinetic spectrum (Lansdall *et al*., 2017). A majority of patients had apathy, which lay near the centre of the multidimensional scaling plot (Figure 3), suggesting that it is related similarly to other features across FTLD syndromes. The behavioural syndrome dimension was expressed across multiple groups and was not restricted to the subset of the cohort with bvFTD (Figure 4A). Interestingly, not all patients with bvFTD had very high scores on this behavioural syndrome dimension. Those with lower behaviour scores, but a clinical diagnosis of bvFTD, may represent bvFTD with prominent apathetic/dysexecutive symptoms (O’Connor *et al*., 2017), or reflect more advanced disease, when many of the more florid behavioural changes are less pronounced (O’Connor *et al*., 2016). A proportion of patients with PSP and CBS had high scores on this syndrome dimension. Behavioural changes in PSP and CBS are well recognised (Burrell *et al*., 2014), but are often thought to be mild. Our findings suggest that behavioural impairments in PSP and CBS can very prominent, in fact some patients with PSP and CBS had higher scores on syndrome dimension one than some with bvFTD. Importantly, no other clinical feature had negative loading coefficients on syndrome dimension one, suggesting that behavioural features can coexist with all other FTLD-related features. Global cognitive impairment was represented by syndrome dimension two. All the Addenbrooke’s Cognitive Examination subscores and carer ratings of everyday skills and memory loaded onto this dimension. However, the reasons for low ACER scores may vary depending on which other symptom profiles are expressed: a low score on the ACER could be due to progressive dementia or caused by severe behavioural (syndrome dimension one) or language (dimension five) or motor (dimensions three and four) impairment, all of which would interfere with the test session.

Our results are also relevant to the current nosology of primary progressive aphasias. Semantic impairments loaded onto a different syndrome dimension and clustered separately from the language impairments associated with non-fluent and logopenic primary progressive aphasia. This provides partial support for the current distinction between svPPA and other forms of PPA. However, nfvPPA and lvPPA were not readily distinguished by the data-driven analysis – as has been noted in a previous independent cohort (Sajjadi *et al*., 2012). In contrast, patients with svPPA were similar to bvFTD in many respects (Figure 4), compulsive behaviours, stereotyped speech and simple repetitive habits were closely linked to semantic language impairments, including object recognition and single word comprehension (Harris *et al*., 2016). Other language features, including impaired syntactic comprehension, agrammatism and speech apraxia, were closely related to CBS-like motor features (syndrome dimension 3), in CBS, PSP, and nfvPPA groups - in keeping with the well characterised overlap of non-fluent (J. D. J. Rohrer *et al*., 2010; J. Rohrer *et al*., 2010) and apraxic (Josephs *et al*., 2006, 2012) speech with PSP and CBS (Armstrong *et al*., 2013; Respondek and Höglinger, 2016; Peterson *et al*., 2019). The PPA diagnostic criteria require that language impairments are the most prominent clinical feature and the principal cause of difficulty with activities of daily living.

This may not be the case in some patients with svPPA; although clinicians may note prominent semantic impairments, co-existent behavioural impairment may be more conspicuous to relatives or carers and have a greater impact on independence and daily living. In addition, we report the practical difficulties applying the current PPA diagnostic criteria. In our epidemiological-based cohort nineteen patients met criteria for primary progressive aphasia (Gorno-Tempini *et al*., 2011) but not one of the PPA subtypes. The current diagnostic criteria are stringent and require the presence and absence of multiple language features. Patients with language symptoms may have very isolated deficits (Josephs *et al*., 2012) or at the other extreme multiple impairments which span more than one PPA subtype, even at diagnosis (Utianski *et al*., 2019).

Many studies have correlated clinical syndromes with structural change, using computational morphometry on volume, thickness, curvature or cortical diffusivity. Typically, these compare patient groups to each other or to controls, to reveal group-based patterns of atrophy in bvFTD (Schroeter *et al*., 2007; Whitwell *et al*., 2012; Kamalini G. Ranasinghe *et al*., 2016; Meeter *et al*., 2017; Perry *et al*., 2017; Y. Chen *et al*., 2018; Illán-Gala *et al*., 2019), svPPA (Gorno-Tempini *et al*., 2004; Schroeter *et al*., 2007; Kumfor *et al*., 2016), nfvPPA (Gorno-Tempini *et al*., 2004; Schroeter *et al*., 2007; Santos-Santos *et al*., 2016), PSP (Brenneis *et al*., 2004; Lagarde *et al*., 2013; Piattella *et al*., 2015; Dutt *et al*., 2016; Whitwell, Höglinger, *et al*., 2017; Whitwell *et al*., 2019) and CBS (Josephs *et al*., 2010; Whitwell *et al*., 2010; Dutt *et al*., 2016). However, these previous methods are limited by the categorical approach to diagnosis. In order to reveal the associations between phenotypic features and structural change, across diagnostic groups, we used source based morphometry to identify regions of covarying atrophy patterns (Xu *et al*., 2009). We confirmed our hypothesis that individual atrophy patterns are not confined to specific diagnostic groups. Our imaging cohort was generally representative of the whole FTLD population, with similar weightings across five out of six dimensions and demographics. Participants who underwent MRI were less affected in the global cognitive impairment syndrome dimension, likely due to the practical difficulties of scanning participants with advanced dementia. Frontal lobe atrophy patterns were seen in participants from all groups, especially bvFTD and PSP. Subcortical atrophy was more prevalent in PSP and CBS but was also seen in bvFTD and PPA, and a majority of bvFTD patients had negative scores on the basal ganglia imaging component. This has been noted previously in symptomatic bvFTD and PPA (Schroeter *et al*., 2007; Bocchetta *et al*., 2018), and those at genetic risk of FTD (Rohrer *et al*., 2015). Brainstem atrophy, while characteristic of PSP (Whitwell, Höglinger, *et al*., 2017), was also seen in some patients with CBS and nfvPPA, but this has previously been shown not to predict PSP pathology (Whitwell *et al*., 2013). The source based morphometry approach also revealed a group of patients who are not well accommodated in the current diagnostic criteria. Five patients with a nominal diagnosis of bvFTD had very low scores on the right temporal lobe imaging component, and we suggest that these might better be called the right variant of semantic dementia, which causes a combination of behavioural and semantic impairments with prosopagnosia (Chan *et al*., 2009; Kumfor *et al*., 2016). A subset of patients with CBS and mixed PPA had negative scores on component 8, indicating posterior cortical atrophy. These patients may be more likely to have Alzheimer’s Disease pathology (Lee *et al*., 2011).

We identified three significant canonical “structure-function” correlations in the cohort (Figure 6). These represent the spectrums of anatomical change underlying behavioural, motor and language impairments. These structure-function correlations did not replicate classical nosological distinctions. Instead they provide an alternative data-driven approach with which to understand and target treatments for syndromes associated with FTLD. The first canonical correlation found an association between motor cortex and brainstem atrophy with PSP or CBS-like motor impairments. Unsurprisingly, PSP and CBS had significant correlations between these canonical covariates but so did bvFTD and nfvPPA, reflecting the motor impairments that are seen in a subgroup of these patients. The second canonical correlation represented the spectrum between frontotemporal (positive scores) and posterior cortical atrophy (negative scores). This canonical covariate may differentiate FTLD from Alzheimer’s disease pathology, as negative scores on this imaging covariate resemble an AD-like atrophy pattern. The third canonical covariate was associated with significant correlations in all FTLD subgroups apart from bvFTD, and encompassed a range of cognitive, behaviour and motor clinical features associated with cortical and subcortical atrophy.

Longitudinal analysis in a subset of patients confirmed that, with disease progression, overlap between FTLD phenotypes increases (Kertesz *et al*., 2005). A greater number of patients met criteria for several FTLD subtypes compared to first assessment and there was greater overlap between all syndrome dimensions (Figure 7). Our transdiagnostic approach allows disease progression to be more accurately represented, in terms of worsening clinical features rather than conflicting diagnoses. Assessing FTLD syndromes in isolation, without reference to the whole FTLD syndrome spectrum, risks missing evolving signs of other FTLD syndromes and therefore underestimating disease severity. The time between the two phenotypic assessments was relatively long (mean 3.6 years) given the mean survival in FTLD syndromes (Coyle-Gilchrist *et al*., 2016); therefore these results may be biased towards patients with more slowly progressive disease.

A strength of our analysis is that it is embedded within an epidemiological cohort study. Previous structure-function studies of these disorders may have been influenced by low sample sizes and selection bias, by focussing only on patients at earlier disease stages who are well enough to attend subspecialist research centres for detailed phenotypic assessment. The representativeness in our study may partly explain why many of our patients lay across diagnostic criteria. However, our study also has several limitations. Applying multiple diagnostic criteria across all patients raises challenges. For example, the criteria can include an exclusion clause, that the illness is not better explained by another diagnosis. We lifted this criterion and applied the clinical features to the other positive and negative criteria. Patients may have symptoms or signs that do not quite reach threshold needed to meet a diagnostic criterion. Our approach was to try to apply the same threshold in all groups, in asserting the presence of a symptom or sign. Our assessment of clinical features was also cross sectional, rather than a retrospective estimate of presenting features. Some of the diagnostic criteria (e.g. for PPA (Gorno-Tempini *et al*., 2011)) refer to the dominance of a symptom cluster (eg language disorder) at presentation. This sounds straightforward, but the time of presentation varies widely, is often late (Coyle-Gilchrist *et al*., 2016), and is partially dependent on variations in healthcare services, referral pathways and public awareness of symptoms’ significance (Bradford *et al*., 2009). These factors interfere with the ability of symptomatology to inform the diagnosis and likely pathology, especially in overlap syndromes such as CBS-NAV, or PSP-F. This transdiagnostic approach to FTLD may not be appropriate in all situations, for example trials of treatments targeting at a specific proteinopathy. Currently there are no robust biomarkers that can differentiate between, for example FTLD-tau and FTLD-TDP43 (Bevan-Jones *et al*., 2017; Meeter *et al*., 2017), and current trials focus recruitment on subsets of patients with strong clinicopathological correlation like PSP-RS (Boxer *et al*., 2019). However, this limits patient access to drug trials, given the poor clinicopathological correlation in the majority of FTLD syndromes. Emergence of more accurate biomarkers, whether PET, CSF or blood based (Meeter *et al*., 2017; Leuzy *et al*., 2019), may allow a more transdiagnostic approach. This would facilitate accurate drug targeting while maximising power and generalisability of results.

Research related to disease nosology often raises the issue of whether to ‘lump’ disorders together or to ‘split’ them into subtypes (Scaravilli *et al*., 2005). There may be occasions where the decision to lump or split aids insight into the neurobiology of disease. But, lumping and splitting can also obscure insights. We propose an alternative approach, with data-driven spectral analyses, that neither lump nor split arbitrarily, but allow phenotypic and imaging variance to elucidate pathogenesis of cognitive syndromes. We acknowledge however that our brain metrics are only crude measures of atrophy. Other brain measures, of tau burden (Passamonti *et al*., 2017; Whitwell, Lowe, *et al*., 2017; Bevan-Jones *et al*., 2019), synaptic density (M. K. Chen *et al*., 2018), physiology (Hughes *et al*., 2018; Sami *et al*., 2018) and functional connectivity (Seeley *et al*., 2009; Rittman *et al*., 2019) may enrich the source based morphometric approach, integrating PET markers of pathology (Passamonti *et al*., 2019) or spectroscopic measures of the neurotransmitter deficits in FTLD (Kantarci *et al*., 2010; Murley and Rowe, 2018). Genetic information could further inform the multivariate analysis of phenotype, mindful that while bvFTD has a strong genetic component, svPPA and PSP do not (Rohrer *et al*., 2009). An additional limitation is the potential for multiple pathologies, in which several pathogenic protein inclusions may co-exist and be synergistic in neurodegeneration (Robinson *et al*., 2018).

In conclusion, we have presented evidence from a transdiagnostic, data-driven approach to the clinical and structural phenotypes in syndromes associated with FTLD. Patient categorisation and selection should depend on the study or question of interest (Husain, 2017; Coulthard and Love, 2018), but for understanding the origin of symptoms, designing symptomatic treatment, and assessment of diagnostic biomarkers, we suggest that the more relevant outcomes are the data-driven axes of disease. Clinical heterogeneity and phenotypic variance are ‘noise’ in category-based analysis of disease and treatment effects and undermine the observation of effects. However, the same variance can be informative in terms of a spectrum of structure-function abnormality, complementing data-driven approaches to order neurodegenerative disease using neuropathological features (Cornblath *et al*., 2019). The adoption of such a data-driven approach provides a comprehensive framework with which to understand disease progression and heterogeneity.

## Data Availability

Anonymised data are available on reasonable request for academic purposes.

## Abbreviations

FTLD: Frontotemporal Lobar Degeneration
bvFTD: Behavioural variant frontotemporal dementia
PPA: Primary progressive aphasia
nfvPPA: Non-fluent variant primary progressive aphasia
svPPA: Semantic variant primary progressive aphasia
lvPPA: Logopenic variant primary progressive aphasia
PSP: Progressive supranuclear palsy
CBS: Corticobasal syndrome
CBD: Corticobasal degeneration
ACER: Addenbrooke’s Cognitive Examination – Revised
CBIR: Cambridge Behavioural Inventory - Revised

## Acknowledgements

We would like to thank the patients and their families and carers, the radiographers at the Wolfson Brain Imaging Centre, University of Cambridge and all the staff at the Cambridge Centre for Frontotemporal Dementia and Related Disorders, University of Cambridge.

## Funding

This work was funded by the Holt Fellowship (AGM), British Academy (KAT, PF160048), Wellcome Trust (JBR, 103838), the PSP Association, the Medical Research Council, the National Institute for Health Research Cambridge Biomedical Research Centre and Cambridge Brain Bank; and the Cambridge Centre for Parkinson Plus.

## Competing Interests

JBR serves as an associate editor to Brain, and is a non-remunerated trustee of the Guarantors of Brain and the PSP Association (UK). He provides consultancy to Asceneuron, Biogen, UCB and has research grants from AZ-Medimmune, Janssen, Lilly as industry partners in the Dementias Platform UK.

